# Associations between Cardiovascular Risk Factors and Neurofilament Light Levels Among U.S. Mexican American Adults

**DOI:** 10.1101/2025.03.13.25323894

**Authors:** Monica M. Diaz, Eran Dayan, the Health and Aging Brain Study: Health Disparities (HABS-HD) Team

**Affiliations:** Department of Neurology, University of North Carolina at Chapel Hill School of Medicine, Chapel Hill, NC USA; Department of Radiology and Biomedical Research Imaging Center, University of North Carolina at Chapel Hill School of Medicine, Chapel Hill, NC USA

**Author notes:** **Corresponding author:** Eran Dayan, PhD, University of North Carolina at Chapel Hill Dept of Radiology, CB# 7510, Chapel Hill, NC 27599-7510, Monica M. Diaz, MD, MS, 170 Manning Drive, CB 7025 Chapel Hill, NC 27599-7510. HABS-HD MPIs: Sid E O’Bryant, Kristine Yaffe, Arthur Toga, Robert Rissman, & Leigh Johnson; and the HABS-HD Investigators: Meredith Braskie, Kevin King, James R Hall, Melissa Petersen, Raymond Palmer, Robert Barber, Yonggang Shi, Fan Zhang, Rajesh Nandy, Roderick McColl, David Mason, Bradley Christian, Nicole Philips and Stephanie Large.

**Keywords:** Neurofilament light protein, cardiovascular disease, white matter hyperintensities, Mexican American, Latinos, healthcare disparities

## Abstract

**Background:** The pathophysiological mechanisms that may differentially impact brain health and cognitive aging outcomes among Latino compared with non-Latino White (NLW) adults in the U.S remain incompletely understood. Recent evidence suggests that neurofilament light (NfL) levels, a biomarker of neuronal injury predictive of dementia risk, is associated with cardiovascular risk factors in both Latino and NLW populations. The current study examines whether associations between plasma NfL levels and markers for cardiovascular health differ among U.S. Mexican American (MA) and NLW adults enrolled in the Health and Aging Brain Study: Health Disparities (HABS-HD).

**Methods:** Data from 1317 participants (648 MA and 669 NLW) were analyzed, including phenotypic, neuroimaging, and plasma NfL data. Cardiovascular health factors included total volume of white matter hyperintensities (WMH), and diagnoses of hypertension, diabetes, and CVD.

**Results:** We found that NfL burden levels among MA and NLW participants differed as a function of diabetes and CVD diagnosis, with steeper differences observed in the MA group. Additionally, the association between WMH volume and NfL varied between the two groups, with a steeper association observed in the MA group.

**Conclusions:** These findings highlight the potential utility of NfL as a prognostic biomarker for CVD and neurodegeneration, particularly among MA adults. Further research is needed to clarify the mechanisms underlying these associations and to develop targeted neurodegenerative prevention strategies that address disparities in brain aging.

## Introduction

Older Latino adults in the U.S are roughly 1.5 times more likely to develop Alzheimer’s disease and related dementias (ADRD) compared to non-Latino White (NLW) adults of similar ages (1). ADRD prevalence is projected to dramatically increase in the next decades due to changes in life expectancy (2). Given the substantial projected growth of the U.S Latino community (3), particularly that of U.S Mexican Americans (MA) (4), the anticipated impact on ADRD prevalence in this population is alarming. It is thus critical to further investigate the pathophysiological mechanisms that may differentially impact ADRD risk among MA adults compared with NLW in the U.S.

A viable strategy for identifying mechanisms that may contribute to differential brain aging outcomes among MA and NLW is via the use of biomarkers. An emerging biomarker for neurodegeneration and axonal injury is the neurofilament light (NfL) protein. Although present in normal aging, NfL levels have also been associated with other neurodegenerative processes, including multiple sclerosis (5,6), Parkinson’s disease (7) and ADRD (8). The literature points to strong associations between NfL and various markers of cardiovascular health. Namely, plasma NfL levels are associated with white matter hyperintensities (WMH) volume (9,10), an incidental magnetic resonance imaging (MRI) finding, widely considered as reflecting small vessel disease (11). Moreover, higher plasma NfL has also been shown to associate with cardiovascular mortality and CVD (12), and with risk for CVD, coronary heart disease, and heart failure (13). Plasma NfL levels among Latinos who identify as MA have been reported to associate with CVD risk factors, including hypertension, dyslipidemia and diabetes (14). However, the extent to which associations between CVD risk factors and NfL levels differ among Latinos, particularly those who identify as MA, and NLW of similar ages remains incompletely understood.

The aim of the present study was to examine if associations between plasma NfL levels and markers for cardiovascular health differ among U.S. MA and NLW adult participants in the Health and Aging Brain Study: Health Disparities (HABS-HD). We considered data from a total of 1317 participants (648 MA and 669 NLW; **Fig. 1A**), including phenotypic, neuroimaging and plasma NfL data (**Fig 2B**). Cardiovascular health factors included total volume of WMH, and participants’ diagnoses of hypertension, diabetes and CVD. We hypothesized that cardiovascular health factors would differentially impact NfL levels in MA and NLW participants (**Fig. 1C**).

**Figure 1.**
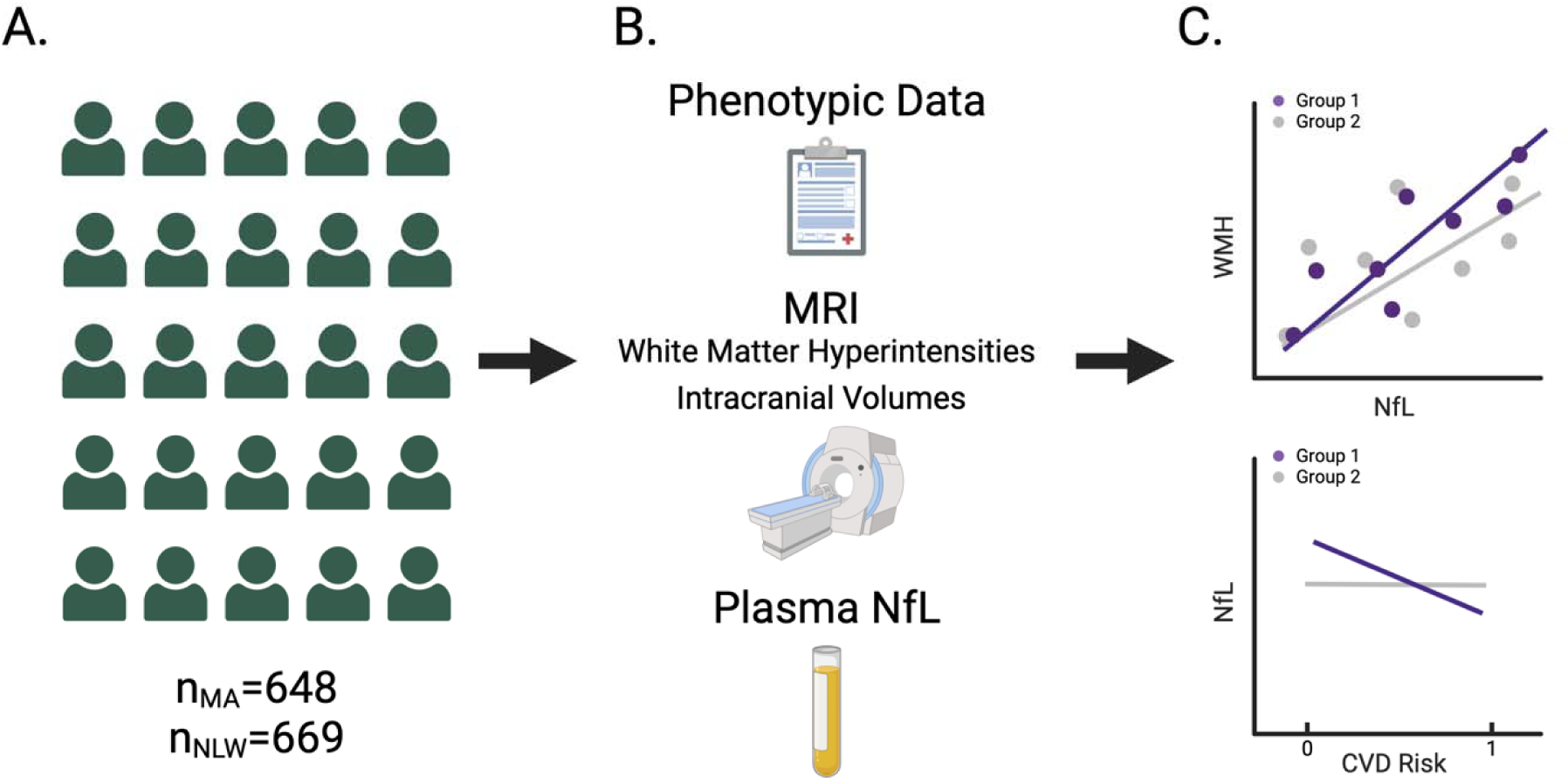
Participants and study design. (A.). Data from a total of 1317 participants were analyzed, including 648 who self-identify as Mexican Americans (MA group), and 669 non-Latino White participants (NLW group). (B). All participants had phenotypic data, MRI data (including white matter hyperintensity and total intracranial volumes) and plasma NfL data. (C.) Analyses focused on associations between NfL burden and WMH volume in the MA and NLW groups, and on the contribution of cardiovascular risk factors to NfL burden in these groups.

**Figure 2.**
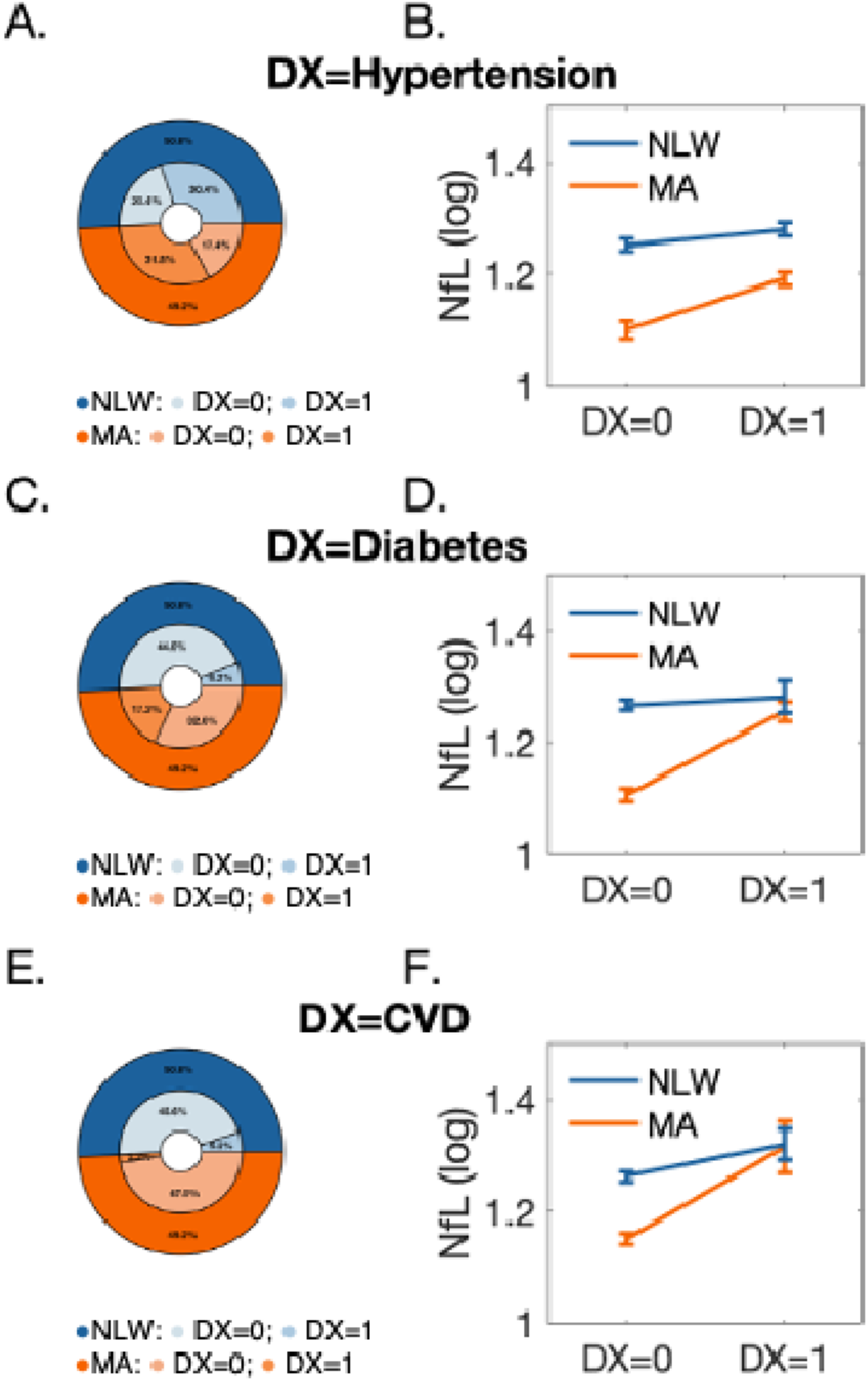
Impact of cardiovascular risk factors on neurofilament light burden. (A). Distribution of hypertension diagnosis did not differ significantly between MA and NLW participants (p=0.068). (B.) NfL burden among individuals in the MA and NLW groups did not differ significantly as a function of hypertension (p= 0.106, for group × diagnosis interaction). (C.) Distributions of diabetes diagnosis differed significantly between the two groups (p<0.001). (D.) NfL burden among individuals in the MA and NLW groups differed significantly as a function of diabetes diagnosis (p< 0.001, for group × diagnosis interaction). (E.) Distributions of cardiovascular disease (CVD) diagnosis were significantly different among the MA and NLW groups (p<0.001). (D.) NfL burden among the two groups differed significantly as a function of CVD diagnosis (p= 0.007, for group × diagnosis interaction).

## Methods

### Participants

All data were collected as part of the HABS-HD (15), a study initiated by the University of North Texas Health Science Center to investigate the factors underlying health disparities in cognitive aging and dementia among MA. Inclusion criteria in the HABS-HD study were self-reported MA or NLW ethnicity, willingness to provide blood samples, ability to undergo neuroimaging, age 50 and older, and fluency in English or Spanish. Exclusion criteria were type 1 diabetes, active infections, current or recent (<12 months) cancer other than skin cancer, current diagnosis of severe mental illness that can impact cognition (excluding depression), recent (<12 months) traumatic brain injury with loss of consciousness, current or recent substance abuse, active medical conditions that can impact cognition, and current diagnosis of non-AD dementia. Here we considered baseline, cross-sectional data from participants who self-reported MA (n=648) and or NLW ethnicity (n=669) who had available neuroimaging (WMH volume and intracranial volume [ICV]), and plasma NfL data. All subjects provided written informed consent, and study procedures were all approved by the local Institutional Review Board.

### Neuroimaging data acquisition and metric extraction

Multimodal magnetic resonance imaging (MRI) data were acquired from participants using a 3T Siemens Magnetom SKYRA scanner. Here, we analyzed data acquired with 3D T1 weighted whole brain magnetization prepared rapid acquisition gradient echo (MPRAGE) (1.1 x 1.1 x 1.2 mm; TR = 2300 ms; TE = 2.93 ms) and fluid attenuated inversion recovery (FLAIR) (1.0 x 1.0 x 1.2 mm; TR = 4800 ms ; TE = 441 ms) sequences. Total volumes of WMH were estimated using FLAIR and MPRAGE data by the HABS-HD team (16) with the Lesion Growth Algorithm(17), available in the Lesion Segmentation Toolbox in the Statistical Parametric Mapping (SPM) software suite. ICVs were estimated for each participant using the segmentation pipeline (18) in FreeSurfer 6.0 and based on MPRAGE data. Total WMH and ICV were both obtained from the HABS-HD dataset as derived variables.

### Blood sample collection and processing

Procedures for blood collection and processing were described elsewhere (14,15). Briefly, fasting blood samples were collected and processed within 2 hours post-collection, in accordance with established guidelines (19). Plasma NfL was assayed from participants’ samples using the Quanterix single molecule array (Simoa HD1) platform.

### Impact of Cardiovascular and related diseases

The existence of multiple CVD risk factors was recorded for each participant via self-report and included in the dataset as binary variables (yes/no). This included: (1) hypertension, (2) diabetes (type 2), and (3) CVD.

### Statistical Analysis

All statistical analyses were performed with JASP 0.16.4 and MATLAB R2023.a. Pairwise differences in demographic, imaging or biomarker data were tested with independent sample t tests or Mann-Whitney U tests (used when Gaussian distributions could not be assumed). Categorical variables were compared with a chi-squared test. Correlations between variables were examined with the Pearson correlation coefficient (r). Continuous variables with non-Gaussian distributions (NfL and WMH) were log-transformed in all correlations analyses. The impact of 3 CVD risk factors we consider here (hypertension, diabetes, diagnosis of CVD) on NfL burden was examined with Analysis of Covariance (ANCOVA) models, where NfL burden served as the dependent factor, and sex, group (MA, NLW) and each CVD risk variable served as factors. The models were further adjusted for age and education. To examine if ethnicity differentially impacted the association between WMH and NfL, we performed an ANCOVA where WMH volume served as the dependent variable, group and sex as factors and NfL as a covariate of interest. The model was further adjusted for age, education and ICV. Continuous variables with non-Gaussian distributions (NfL and WMH volume) were log-transformed in the ANCOVA model.

## Results

### Demographics

Data from a total of 1317 participants were analyzed (**Figure 1**), including 648 who self-identified as MA and 669 who identified as NLW. Male-to-female distributions differed between the two groups (χ^2^=23.776, p < .001), with a larger proportion of males in the MA group. Participants in the MA group were also younger (t_1315_=12.755, p < .001), had lower educational attainment (t_1315_=28.621, p < .001), lower total ICV volumes (t_1315_=12.118, p < .001), lower WMH volumes (Mann-Whitney U test, p < .001) and lower NfL burden (Mann-Whitney U test, p < .001).

### Impact of cardiovascular health risk factors on NfL burden

We first examined whether ethnicity interacted with cardiovascular health risk factors and their impact on NfL burden. This analysis considered 3 common cardiovascular health risk factors: hypertension, diabetes, and diagnosis of CVD. Rates of hypertension in the sample analyzed here did not differ between the MA and NLW groups (χ^2^=3.32, p=0.068) (**Fig. 2A**). Moreover, NfL burden in MA and NLW participants did not differ significantly in individuals with and without a diagnosis of hypertension (group × diagnosis interaction: F_1,1310_=2.61, p=.106) (**Fig. 2B**, Supplemental Table 1). Different results were observed for diabetes. Rates of diabetes diagnosis differed among the two groups (χ^2^=96.538, p=< .001) (**Fig. 2C**), with higher rates seen among MA participants. Moreover, NfL burden levels among MA and NLW participants differed as a function of diabetes diagnosis (group × diagnosis interaction: F_1,1310_=14.692, p=< .001). Namely, differences among participants with and without diabetes, were steeper in the MA group (**Fig. 2D**, Supplemental Table 2). Diagnosis of CVD also differed between the two groups (χ^2^=15.299, p=<.001), with higher rates seen in the NLW group (**Fig. 2E**). However, as in diabetes, differences among individuals with and without CVD diagnosis were steeper in the MA group (group× diagnosis interaction: F_1,1310_=7.211, p= .007) (**Fig. 2F**, Supplemental Table 3).

### Impact of ethnicity on association between WMH volume and NfL

We next set out to examine if NfL levels among MA and NLW participants were differentially associated with WMH volume, a widely used marker for small vessel disease in the brain. First, we estimated associations between WMH volume, NfL and the background demographic variables considered here (age and education). Associations between each variable and total ICV were examined as well, since WMH volume is likely to covary with this variable. In all analyses NfL and WMH volume were log-transformed to adjust for their skewed distributions. When considering the entire sample (**Fig. 3A**), strong correlations were observed between NfL and WMH volume (r=0.416, p < .001), and between these two variables and age, education and total ICV (all p values <=.04) . Correlations were generally similar when data from each group was considered separately (**Fig. 3B, 3C**), with a higher correlation observed between WMH and NfL in the MA group (r=0.422, p < .001), compared to the NLW group (r=0.351, p < .001). We thus next compared the strength of these association more directly with an ANCOVA model. This analysis revealed that the association between WMH volume and NfL varied between the two groups (group× NfL interaction: F_1,1298_=8.791, p= .003), with a steeper association observed in the MA group (**Fig 3D**, Supplemental Table 4).

**Figure 3.**
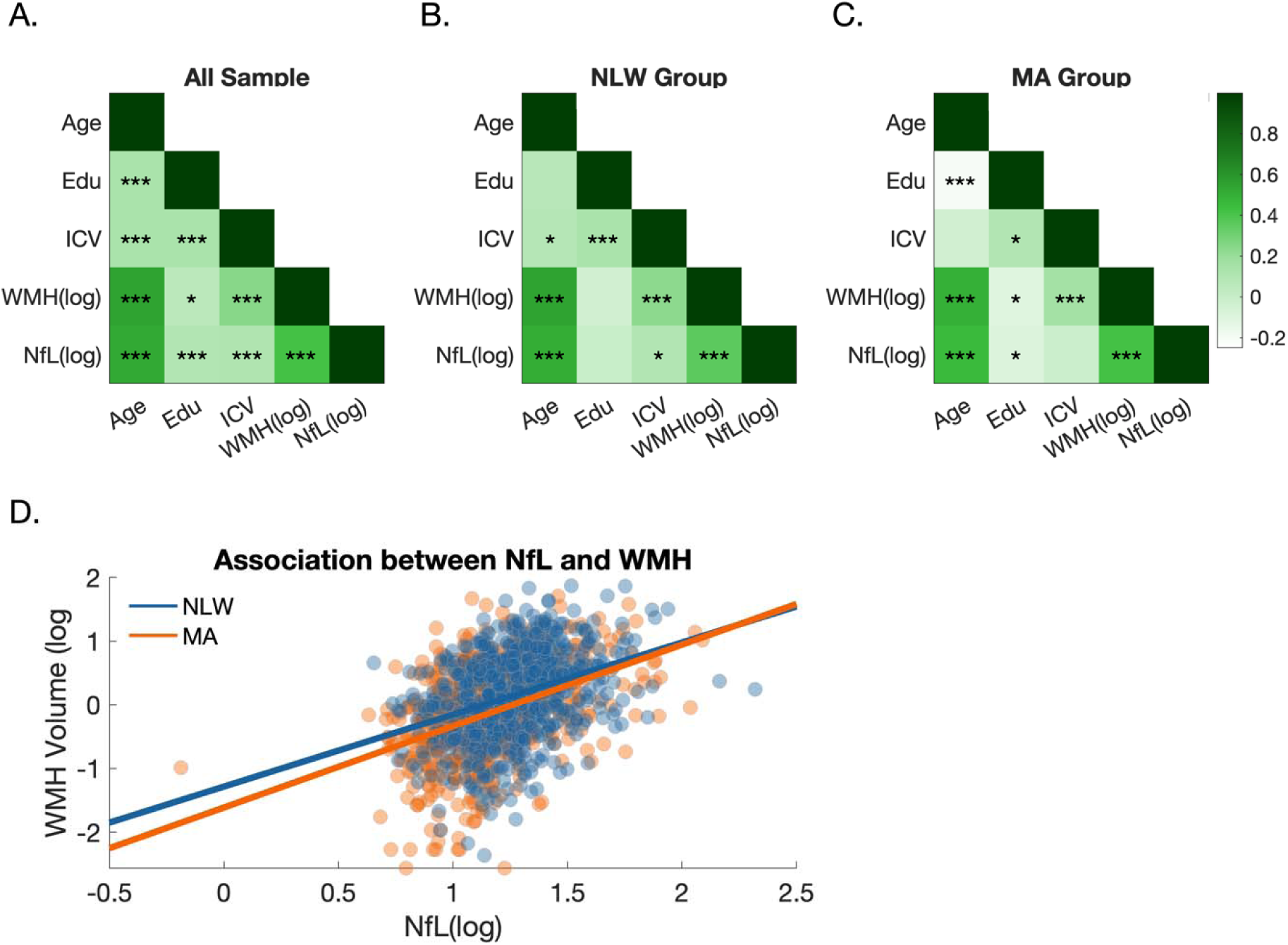
Associations between Neurofilament light burden and White Matter Hyperintensity Volume among U.S. non-Latino Whites and Mexican Americans. (A). Associations (Pearson’s correlations) between age, education, ICV, total WMH volume (log-transformed) and NfL burden (log-transformed) are shown for the entire sample and for the NLW (B.) and MA (C.) groups separately. (D.) The association between NfL burden (log-transformed) and WMH volume (log-transformed) differed significantly between individuals in the MA and NLW groups (p=0.003, for group × NfL interaction, adjusting for age, sex, ICV and education), with a steeper association observed for individuals in the MA group. *p<0.05, ** p<0.01, *** p<0.001

## Discussion

The study examined the potential impact of ethnoracial background on associations between NfL levels and cardiovascular health factors among adults enrolled in the HABS-HD cohort. We found that the existence of cardiovascular health risk factors (particularly diagnosis of diabetes and CVD) impacted NfL levels differently when comparing MA and NLW participants, with a larger impact seen among the MA group. We also found differential associations between WMH volume and plasma NfL levels among MA and NLW participants, wherein the MA group displayed a steeper association than the NLW group.

The association between WMH volume and serum NfL has been demonstrated in previous studies, but the effect of ethnoracial background (comparing Latinos vs. NLW) on this relationship has not been studied. One prior study found that both NfL and WMH volume were significantly independently associated with cognitive decline, with larger WMH volumes influencing cognitive decline rates the most among those with lower NfL levels (20). Another study found an association between higher plasma NfL and WMH volume, but not with global cognitive impairment (9), suggesting that NfL may be a marker of WMH volume but not necessarily cognitive impairment. Similar relationships have been investigated among other ethnoracial groups, such as in Asian cohorts, where high plasma NfL was associated with increased cerebrovascular disease risk and progression (10,21,22). However, some studies did not find significant relationships between WMH and plasma NfL (23), highlighting the need for further investigation in larger cohorts and across different ethnoracial backgrounds.

Our study is, to the best of our knowledge, the first to study the relationship between NfL levels and WMH volumes as a function of ethnoracial background, specifically comparing MA and NLW adults. The HABS-HD group found that a higher Framingham Risk Score (indicating higher CVD burden) was associated with higher NfL levels only among MA adults and not among NLW. However, this relationship with WMH volume was not explored. Similar to our study, hypertension was associated with NfL in both groups (24), but the degree of the association on MA compared with NLW was not studied at the time.

Our results identify relationships between CVD and NfL, as well as diabetes and NfL, with a stronger impact among MA compared to NLW. Previous studies have demonstrated the association between CVD, including hypertension and cardiovascular death and stroke/transient ischemic attack, and higher serum NfL(25–27), suggesting its utility as a predictive marker for cardiovascular outcomes. Our findings indicate that these associations may be stronger for MA, highlighting the potential utility of using NfL as a prognostic biomarker for CVD in this population.

This study has several limitations worth noting. Firstly, it focuses on a U.S. MA cohort, which restricts the applicability of our findings to other Latino groups. Secondly, the cross-sectional observational design prevents us from identifying causal relationships. Additional factors related to WMH volume and NfL burden, such as other cardiovascular risk factors or social and cultural factors, may partly explain the observed differences in white matter changes and NfL burden between MA and NLW groups. Additionally, our study did not consider cognitive impairment or decline as an outcome, which has been shown to differentially associate with WMH volume among MA and NLW (29). Whether the associations reported here between NfL and cardiovascular health further impact cognitive outcomes should be explored in larger studies, along with mechanisms of reserve and resilience (30,31) that may mitigate detrimental impact on cognitive outcomes. It is also important to mention that many white matter diseases (32) may not be fully captured via quantification of metrics such as WMH volume. Additional indices capturing white matter disease should be integrated in future research focusing on more diverse study populations.

In summary, our findings indicate that the relationship between NfL burden and markers for cardiovascular health varies across ethnoracial groups, with a stronger association observed among MA compared to NLW. These results have significant clinical implications. Specifically, for patients diagnosed with cerebral white matter disease, encouraging modifiable lifestyle changes may enhance cardiovascular health and subsequently reduce neurodegeneration, particularly in MA. Further research involving diverse Latino samples, interventional designs, longitudinal observations, comprehensive examination of cardiovascular health and sociocultural risk factors and use of other white matter indices is necessary to deepen our understanding of the connections between WMH, cardiovascular health risk factors and neurodegeneration in MA and other Latino groups at high risk for dementia.

**Table 1.** Descriptive and inferential statistics of demographics and study outcomes in the study.

|  | MA (n=648) |  | NLW (n=669) |  | *t* | *p* | *d* |
| --- | --- | --- | --- | --- | --- | --- | --- |
|  | *M* | *SD* | *M* | *SD* |  |  |  |
| Demographic Variables |  |  |  |  |  |  |  |
| Age | 63.30 | 8.10 | 69.22 | 8.72 | 12.76 | <.001 | 0.703 |
| Education (years) | 9.63 | 4.65 | 15.50 | 2.52 | 28.62 | <.001 | 1.578 |
| Study Outcomes |  |  |  |  |  |  |  |
| ICV | 1.35×10+6 | 156699.76 | 1.46×10+6 | 187338.16 | 12.12 | <.001 | 0.668 |
| WMH volume | 2.44 | 4.94 | 4.05 | 7.46 |  | <.001 |  |
| NfL | 17.25 | 13.18 | 21.23 | 14.10 |  | <.001 |  |

Note: MA = Mexican American; NLW = non-Latino White; MRI = magnetic resonance imaging; WMH = white matter hyperintensity; ICV = intracranial volume; NFL = neurofilament light

## Supporting information

Diaz-SupplementaryMaterials

## Acknowledgements

Research reported in this publication was supported by the National Institute on Aging of the National Institutes of Health under Award Numbers R01AG054073, R01AG058533, P41EB015922 and U19AG078109. The content is solely the responsibility of the authors and does not necessarily represent the official views of the National Institutes of Health. The data supporting the findings of this study are openly available through the Health and Aging Brain Study: Health Disparities (HABS-HD). Data requests can be made here: https://apps.unthsc.edu/itr/studies/habs.

## Competing Interests and Sources of Financial Support (Including Grant Numbers)

The authors have no conflicts to report. MMD is supported by the NIH NIMH (K23MH131466), Alzheimer’s Association (AARGD-22-924896).

## Data Availability Statement

All data are available online at: https://apps.unthsc.edu/itr/habs-hd

