## Supplementary material for "Associations between Cardiovascular Risk Factors and Neurofilament Light Levels Among U.S. Mexican American Adults": Diaz-SupplementaryMaterials

**Affiliations:**

Supplemental Table 1. *Inferential statistics associated with ethnoracial background* × *hypertension interaction effects on individual neurofilament light levels*

| Variables | *F* | df | *p* |
| --- | --- | --- | --- |
| Ethnicity × Hypertension diagnosis | 2.61 | 1 | .106 |
| Main Effects |  |  |  |
| Hypertension diagnosis | 2.57 | 1 | .109 |
| Age | 358.87 | 1 | <.001 |
| Educational level | 0.44 | 1 | .508 |
| Sex | 0.007 | 1 | .934 |
| Ethnicity | 8.7 | 1 | .003 |
| Residuals |  | 1310 |  |

Supplemental Table 2. *Inferential statistics associated with ethnoracial background* × *diabetes interaction effects on individual neurofilament light levels*

| Variables | *F* | df | *p* |
| --- | --- | --- | --- |
| Ethnicity × Diabetes diagnosis | 14.69 | 1 | <.001 |
| Main Effects |  |  |  |
| Diabetes diagnosis | 22.52 | 1 | <.001 |
| Age | 377.21 | 1 | <.001 |
| Educational level | 0.02 | 1 | .877 |
| Sex | 0.08 | 1 | .783 |
| Ethnicity | 1.27 | 1 | .261 |
| Residuals |  | 1310 |  |

Supplemental Table 3. *Inferential statistics associated with ethnoracial background* × *cardiovascular disease interaction effects on individual neurofilament light levels*

| Variables | *F* | df | *p* |
| --- | --- | --- | --- |
| Ethnicity × CVD diagnosis | 7.21 | 1 | .007 |
| Main Effects |  |  |  |
| CVD diagnosis | 6.66 | 1 | .010 |
| Age | 371.92 | 1 | <.001 |
| Educational level | 1.06 | 1 | .304 |
| Sex | .003 | 1 | .960 |
| Ethnicity | .290 | 1 | .590 |
| Residuals |  | 1310 |  |

Supplemental Table 4. *Inferential statistics associated with ethnoracial background* × *WMH burden interaction effects on individual neurofilament light levels*

| Variables | *F* | df | *p* |
| --- | --- | --- | --- |
| MA ethnicity × WMH volume | 8.77 | 1 | .003 |
| Main Effects |  |  |  |
| Age | 184.36 | 1 | <.001 |
| Educational level | .168 | 1 | .682 |
| Sex | .226 | 1 | .635 |
| ICV | .022 | 1 | .881 |
| WMH volume | 42.34 | 1 | <.001 |
| Ethnicity | 5.23 | 1 | .022 |
| Residuals |  | 1298 |  |
